# Restricted Fluids and Early Vasopressors versus Liberal Fluids and Later Vasopressors for Initial Resuscitation of Septic Shock: Protocol for a Systematic Review and Meta-Analysis

**DOI:** 10.1101/2025.08.28.25334538

**Authors:** Peter G Jones, Sandra L Peake, Alisa M Higgins, Stephen Macdonald, Ary Serpa-Neto, Elissa M Milford, Lorena Romero, Patricia Williams, Conor Keely, Anthony Delaney

**Affiliations:** Adult Emergency Department Auckland City Hospital, Park Road, Grafton Auckland 1172, New Zealand; Department of Surgery, University of Auckland, Park Road, Grafton Auckland 1172, New Zealand; Department of Intensive Care Medicine, The Queen Elizabeth Hospital, Adelaide, South Australia; Australian and New Zealand Intensive Care Research Centre, School of Public Health and Preventive Medicine, Monash University; Faculty of Health and Medical Sciences, University of Adelaide, Adelaide, South Australia; Critical Care Program, The George Institute for Global Health, University of New South Wales, Australia; Centre for Clinical Research in Emergency Medicine, Harry Perkins Institute of Medical Research, University of Western Australia; Department of Intensive Care, Austin Hospital, Melbourne, Australia; Department of Critical Care, Melbourne Medical School, University of Melbourne, Austin Hospital, Melbourne, Australia; Department of Critical Care Medicine, Hospital Israelita Albert Einstein, Sao Paulo, Brazil; Intensive Care Services, Royal Brisbane and Women’s Hospital, Brisbane, Australia; Faculty of Health, Medicine and Behavioural Sciences, The University of Queensland, Brisbane, Australia; Department of Acute and Critical Care, Monash University, Melbourne, Australia; The Ian Potter Library, Alfred Health, Melbourne, Australia; Division of Medicine, Northern Adelaide Local Health Network, Adelaide, South Australia; Malcolm Fisher Department of Intensive Care Medicine, Royal North Shore Hospital, St. Leonards, New South Wales, Australia

## Abstract

**Introduction:** There is uncertainty around optimal haemodynamic management of community acquired septic shock presenting to the Emergency Department (ED), with current guidelines based on low quality evidence. Prior reviews have included mostly patients with hospital-acquired sepsis in the ICU who have received significant fluid resuscitation prior to randomisation. This review will assess whether restricted fluid administration with earlier vasopressor introduction, compared to liberal fluid administration with the potential for later vasopressor introduction, is associated with reduced mortality and improvements in other outcomes in ED patients with early septic shock.

**Methods:** Systematic review and meta-analysis of randomised controlled trials, conducted according to the PRISMA guidelines. There will be no restriction on language, year of publication or publication status. The population will be adult patients predominantly presenting to ED with septic shock. The intervention will be early vasopressor / restricted fluid resuscitation compared to more liberal fluid resuscitation / later vasopressors. The primary outcome is all cause 90-day mortality, with secondary outcomes related to adverse events, ICU interventions, functional status and quality of life up to six months. We will conduct subgroup analyses by pre-randomisation fluid volume and whether timing of initial vasopressor use was pre-specified. We will search major medical databases using structured searches. Studies will be screened for inclusion by two reviewers independently. Data will be extracted onto standardised forms by two reviewers independently. Differences on inclusion and data extraction will be resolved by consensus. A Bayesian framework will be used as the primary statistical approach, and a frequentist framework as the secondary approach. A random-effect model will be used in the analyses and pooled estimates of effect sizes as risk ratios (RRs) for binary outcomes, and mean differences for continuous outcomes will be presented. We will assess risk of bias using the Cochrane Risk of Bias tool (RoB-2) and assess for publication and incomplete outcome reporting. Certainty of evidence will be assessed using the GRADE approach.

**Results:** We will present the results of the review at national and international scientific meetings, and we will prepare a manuscript for submission for publication in a peer reviewed journal.

**Version (date):** V 7.3 (8/8/25)

**Registration:** The protocol was registered with the prospective register of systematic reviews (PROSPERO) and published online on the pre-print server MedRxiv.org on 27th August 2025.

**Contributions:** PJ is the guarantor. PJ, AD, AH, SP and SM drafted the manuscript. All authors contributed to the development of the selection criteria, the risk of bias assessment strategy and data extraction criteria. LR, PJ, AH and AD developed the search strategy. AS provided statistical expertise. All authors read, provided feedback and approved the final version of the protocol.

**Support:** No specific funding was received for this project. Alisa Higgins is supported by an NHMRC investigator grant.

## Introduction

### Rationale

Sepsis is defined as life-threatening organ dysfunction due to a dysregulated host response to infection.^1^ Septic shock is a severe subtype of sepsis characterised by profound abnormalities in perfusion, metabolism and cellular function such that the mean arterial blood pressure (BP) cannot be maintained above 65 mmHg and the serum lactate level is greater than 2 mmol/L.^1^ Globally, sepsis accounts for a high burden of morbidity and mortality.^2^

Patients with sepsis and septic shock often present with hypotension, but there is uncertainty around the best approach to initial haemodynamic resuscitation. In particular, the optimal volume of resuscitation fluid and the timing of introducing vasopressors remain uncertain. The 2021 consensus guidelines from the ‘Surviving Sepsis Campaign’ suggest that at least 30 mL/kg of intravenous (IV) crystalloid fluid be given within the first three hours of resuscitation, although this is based on low quality evidence.^3^ There is no recommendation on when vasopressors should be started in relation to the initial fluid load.^3^

Several large randomised clinical trials have addressed this question. Two of these trials have now been published^4,5^ along with an update of a prior systematic review which included a trial sequential analysis.^6^ Although no difference in mortality was found in the most recent meta-analysis, there was still imprecision around the point estimate of effect, which did not exclude clinically important benefit or harm.^6^ It is important to note that the existing evidence includes data from trials that recruited participants with both community-acquired sepsis (those predominantly presenting to the Emergency Department) and hospital-acquired sepsis. Patients with community acquired sepsis have a different mortality risk profile to hospital-acquired sepsis^7^, and less exposure to intravenous fluids than hospitalised patients. They may also respond differently to fluid therapy and vasopressor strategies compared to patients who develop sepsis during hospitalisation.

## Objectives

This study aims to assess whether restricted fluid administration with earlier vasopressor introduction, compared to liberal fluid administration with the potential for later vasopressor introduction, is associated with reduced mortality and improvements in other outcomes in Emergency Department patients with early septic shock.

## Methods

This systematic review and meta-analysis will be undertaken in accordance with the advice provided by the Cochrane Collaboration^8^ and will be reported in accordance with the Preferred Reporting Items for Systematic Reviews and Meta-analysis statement.^9^ This protocol has been prepared in alignment with the Preferred Reporting Items for Systematic Reviews and Meta-Analysis Protocols 2015 statement^10^

### Eligibility Criteria

#### Study types

We will include randomised clinical trials. There will be no restriction on publication status, language of publication nor year of publication. We will include reports of trials published as full manuscripts or as abstracts where data is available to extract for analysis.

#### Population

We will include trials where the participants are human adults, predominately recruited in the Emergency Department with sepsis associated hypotension or septic shock.

#### Intervention

We will include trials where the intervention is a resuscitation strategy based on the administration of a restricted volume of intravenous fluid with the earlier introduction of vasopressor medications if required to achieve the initial haemodynamic targets.

#### Comparison

We will include trials where the intervention is compared to a strategy of more liberal fluid administration with later vasopressor administration to achieve the initial haemodynamic targets.

#### Outcomes

We will include trials that are able to provide data on any of the outcomes specified below.

We will include trials regardless of the volume of intravenous fluids administered prior to randomisation and regardless of the specific haemodynamic parameters specified in the original trials. We will also include subgroups of patients identified and recruited in the Emergency Department if these subgroups are clearly identified at baseline in trials including broader populations recruited from other settings (e.g., intensive care units or operating theatres). We will exclude studies in paediatric populations (as defined in the source studies) and those studies that predominantly recruited patients with hospital-acquired sepsis. We will exclude studies where patients were randomised to use of a tool to assess fluid responsiveness rather than a particular fluid regimen.

### Information sources

We will search electronic databases MEDLINE (OVID), Embase (Ovid), Web of Science and the Cochrane Central Register of Clinical Trials (OVID), to identify published trials and conference abstracts. We will search clinical trial registries including the World Health Organization International Clinical Trials Registry Platform and Clinicaltrials.gov to identify ongoing or recently completed but unpublished trials. We will include PubMed indexed pre-print servers to identify unpublished studies and will contact experts in the field. We will manually search the reference lists of relevant trials and systematic reviews identified in our search.

### Search Strategy

We developed our search strategy in collaboration with a research librarian (LR). We used a combination of terms for “sepsis and septic shock”, “intravenous fluid therapy” and “vasopressors”, and “emergency department or emergency medicine” combined with sensitive filters to identify randomised clinical trials specific to each electronic database. The full details of the search strategy are included in the Appendix.

### Study records

#### Data management

Study records identified by the search will be loaded into Covidence systematic review software, Veritas Health Innovation, Melbourne, Australia (available at www.covidence.org). Duplicate records will be identified and deleted.

### Selection process

Two authors will independently screen all titles and abstracts to identify potentially eligible studies, with conflicts resolved by a third reviewer. Two authors will independently review the full text reports to assess if they meet all eligibility criteria. Disagreements will be resolved by consensus or by recourse to a third reviewer.

### Data collection process

Data extraction forms will be developed and piloted prior to data collection. Data will be extracted in duplicate with discrepancies resolved by discussion or review by an independent reviewer. Authors of included studies will be contacted twice by email to clarify any uncertainty or to provide missing data.

### Data items

We will collect data from the included trials regarding the study characteristics (first author, year of publication, study period, recruiting countries, number of centres, number of trial participants, study setting). We will also collect data regarding the included populations at baseline (age and sex distribution, severity of illness, lactate, BP, pre-randomisation fluid therapy, pre-randomisation vasopressor therapy, antibiotic administration, source of infection), and details of study interventions (time period, fluid strategy, fluid administered at 6 and 24 hours, receipt of vasopressors at 6 and 24 hours, fluid balance at 6 and 24 hours)

### Outcomes and prioritisation

#### Primary Outcome

The primary outcome will be 90-day all-cause mortality. For trials that do not report 90-day mortality, the nearest available time point (before or after 90 days) will be used.

#### Subgroups for the primary outcome

We will examine the effect of the intervention compared to control in two pre-specified subgroups

- Pre-enrolment fluid volume

- We hypothesise that the estimated treatment effect will be greater in trials with a more restrictive approach to fluid administration prior to randomisation. We will group studies by pre-randomisation fluid volume given (≤1.5L and >1.5L).
- Administration of vasopressors

- We hypothesise that the treatment effect will be greater in trials that specify early use of vasopressors in the restrictive fluid strategy group relative to trials that only specify fluid restriction

The credibility of any observed subgroup effects will be assessed using the Instrument to assess the Credibility of Effect Modification Analyses (ICEMAN) in randomised controlled trials and meta-analyses.

#### Secondary outcomes

We will also collect data to assess the effect of the intervention strategy compared to control on the following secondary outcomes

1. All-cause mortality at the longest time point reported
2. Days alive and out of hospital at 90-days
3. Receipt and duration of vasopressor therapy
4. Receipt and duration of invasive mechanical ventilation
5. Receipt and duration of acute renal replacement therapy
6. Intensive care length of stay
7. Hospital length of stay
8. Adverse events (as reported in the included trials)
9. Quality of life up to 6 months

### Risk of bias in individual studies

Two authors will independently assess risk of bias for the included studies using the Cochrane risk of Bias tool (RoB-2). To assess risk of bias, all available information will be used, including published protocols, trial registration, statistical analysis plans or conference abstracts. Disagreements between reviewers will be resolved by discussion or by consultation with a third reviewer.

### Data synthesis

A Bayesian framework will be used as the primary statistical approach, and a frequentist framework as the secondary approach. A random-effect model will be used in the analyses and pooled estimates of effect sizes as risk ratios (RRs) for binary outcomes, and mean differences for continuous outcomes will be presented. Continuous variables presented in formats not readily amenable to pooling will be converted to mean and standard deviation (SD) with the method described elsewhere.^11^ Along with the pooled estimates of effect sizes, 95% credible intervals (CrI) for the Bayesian meta-analysis and 95% confidence intervals (CI) for the frequentist model will be presented. 95% CrI will be calculated using the shortest interval method, which for unimodal posteriors is equivalent to the highest posterior density region method. For all analyses, Bayes factors will be based on marginal likelihoods. Trials with zero events (if any) will be included in the final model and an effect estimate calculated accordingly. For the frequentist analysis, a random-effect model using DerSimonian-Laird estimates of the between-study variance will be used.

All analyses will be performed considering a minimally informative (unit information prior for the log-RR) distribution for the effect prior, and a weakly informative half-normal prior distribution with scale 0.5 for the heterogeneity prior. Priors were selected on the basis of previous recommendations.^12^ In sensitivity analyses, the treatment effect prior probability distribution will be defined by setting an optimistic, and a pessimistic prior belief for the treatment effect.^13^ The strength of these prior beliefs (the variance setting to establish the shape of the distribution) will be set as moderate for the optimistic and minimally informative priors and weak for the pessimistic prior. In other words, the priors will be set so that we cannot rule out an eventual benefit but can mostly rule out large effect sizes for the intervention and acknowledge a non-negligible chance of the intervention being harmful. In mathematical form, the minimally informative prior is normally distributed and centred at the absence of effect [OR = 1; log(OR) = 0] with a SD of 0.355, such that 0.95 of the probability falls in the range of 0.5–2. The pessimistic and optimistic priors will be informed by the range of effect size estimates from previous studies assessing the effect of a restrictive resuscitation strategy ranging from a reduction of 5% in mortality to an increase of 2%^6^ (OR = 0.375 for the optimistic prior and OR = 1.33 for the pessimistic prior). The optimistic prior SD will be defined to retain a 0.15 probability of harm [Pr(OR > 1)] (SD = 0.10), and the pessimistic prior will be defined to retain a 0.30 probability of harm [Pr(OR < 1)] (SD = 0.06).^14^ The density distribution of the priors is shown in the Appendix.

Quantitative heterogeneity will be assessed with the posterior estimates of the heterogeneity parameter (tau) with its 95%CrI. The proportion of variation across studies owing to heterogeneity rather than chance will be assessed with the I^2^ statistic. Subgroup analyses, based on the pre-specified subgroups as noted above will be performed. Subgroup heterogeneity will be assessed by including an interaction term in the Bayesian analysis to obtain an estimate and 95% CrI for the ratio of RRs (RRRs) from the posterior distribution of the interaction estimate.

We will also undertake a trial sequential analysis to determine if the available evidence is sufficient to draw a reliable conclusion or whether further evidence is required.

All statistical analyses will be performed with R version 4.3.3 (R Foundation for Statistical Computing) using the *bayesmeta* and *meta* packages.

### Meta-bias(es)

We will evaluate the potential for publication bias and small study effects by visual inspection of funnel plots. We will conduct a formal evaluation with Eggers test if more than 10 trials are included in the quantitative synthesis of the primary outcome. We will also assess whether there is incomplete outcome reporting within studies.

### Confidence in cumulative evidence

We will use the Grading of Recommendations Assessment, Development and Evaluation approach to assess the overall certainty of evidence.^15^ A Summary of Findings Table will be prepared, including the plain language summary based upon the GRADE recommendations. The certainty of evidence will be evaluated in discussion between the authors. For each outcome we will assess the certainty of evidence as high, moderate, low or very low.

#### Outcome and reporting

We will present the results of the review at national and international scientific meetings, and we will prepare a manuscript for submission for publication in a peer reviewed journal. The authorship of the manuscript will be in accordance with guidelines described by the International Committee of Medical Journal Editors.^16^

## Data Availability

This is a protocol for a systematic review, there is no data yet. When there is data it will be available in the published manuscript

## Appendix

### Final searches on four databases

Database(s): **Ovid MEDLINE(R)**

Search Strategy:

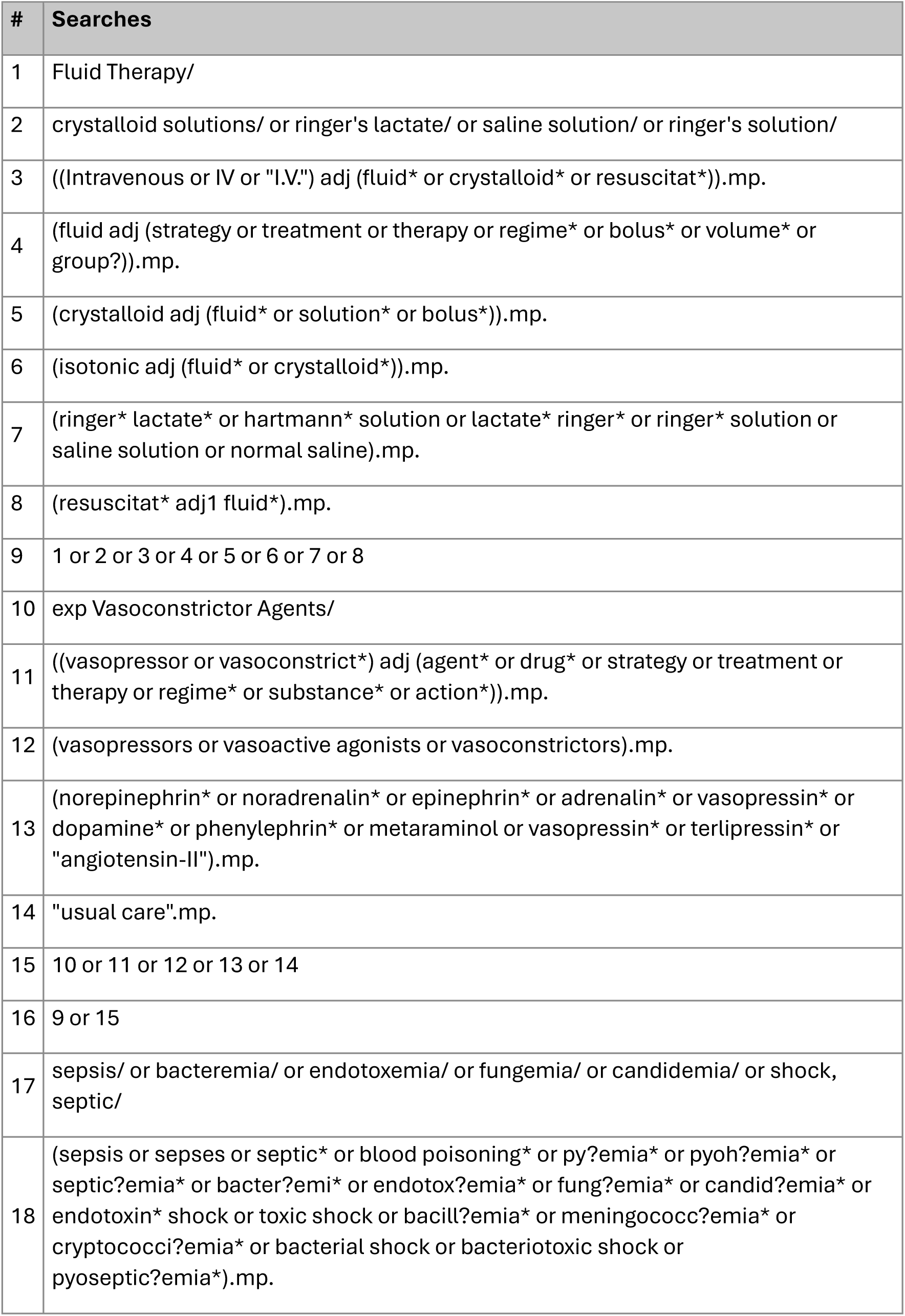
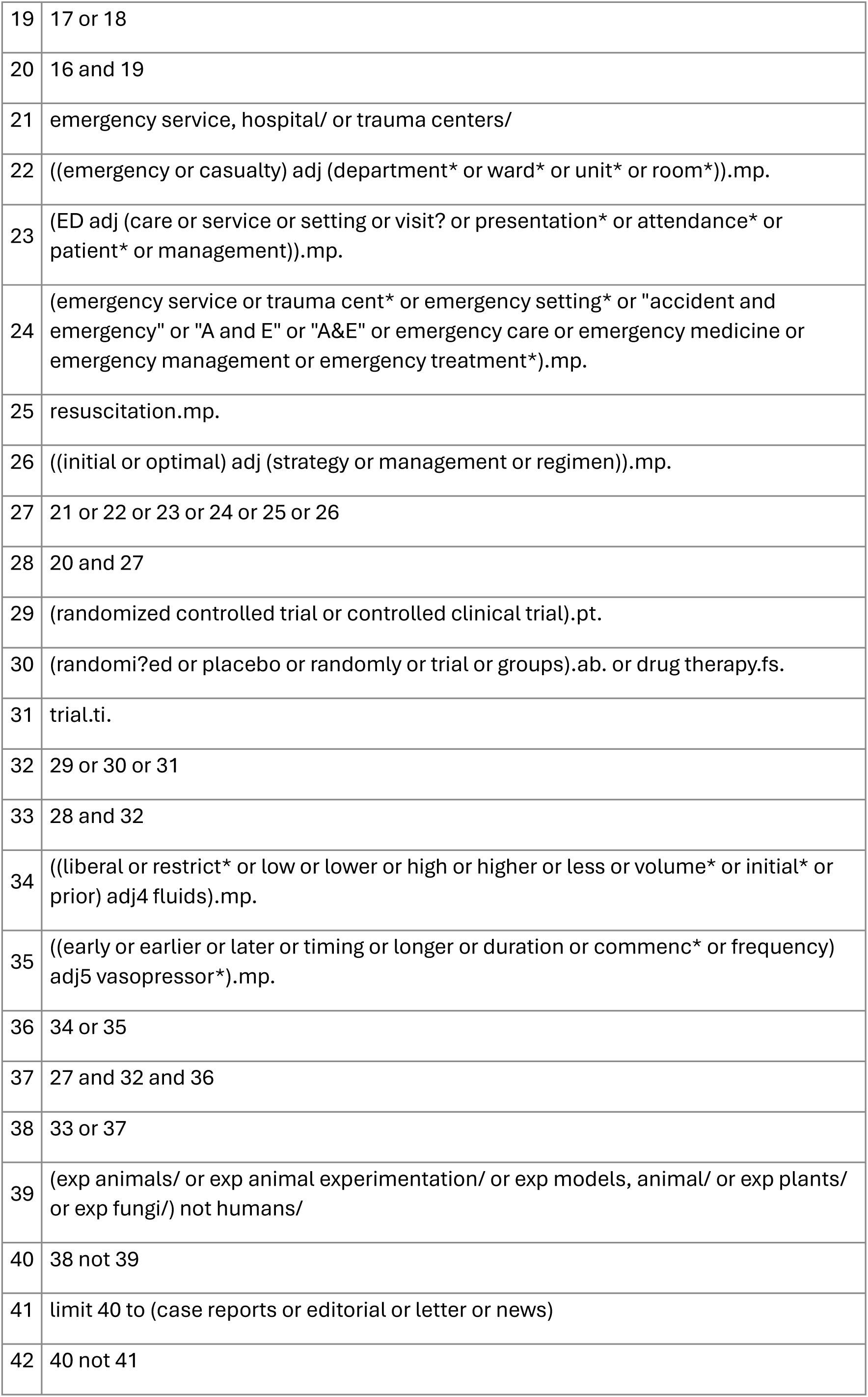

Database(s): **Embase** Search Strategy:

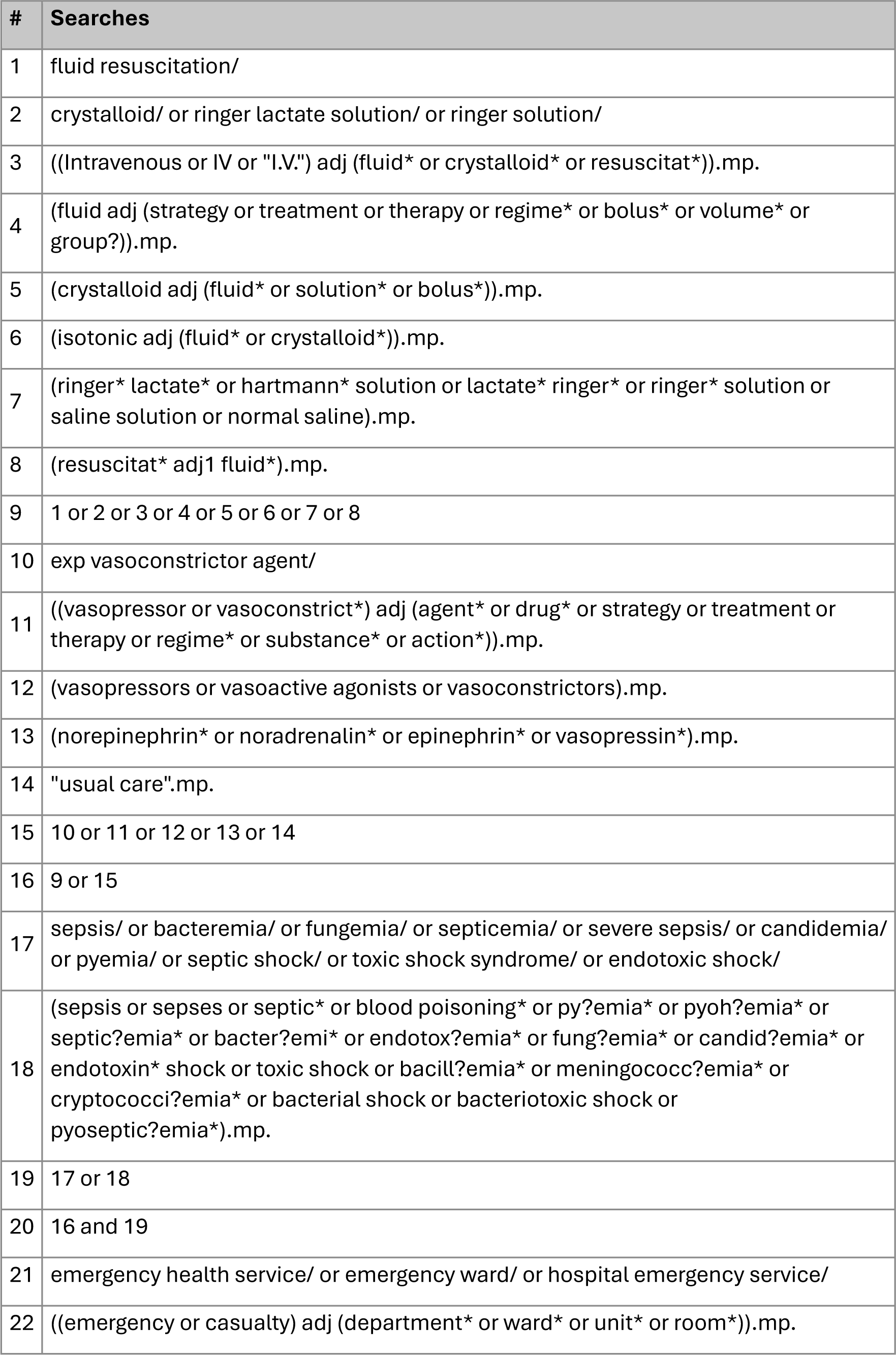
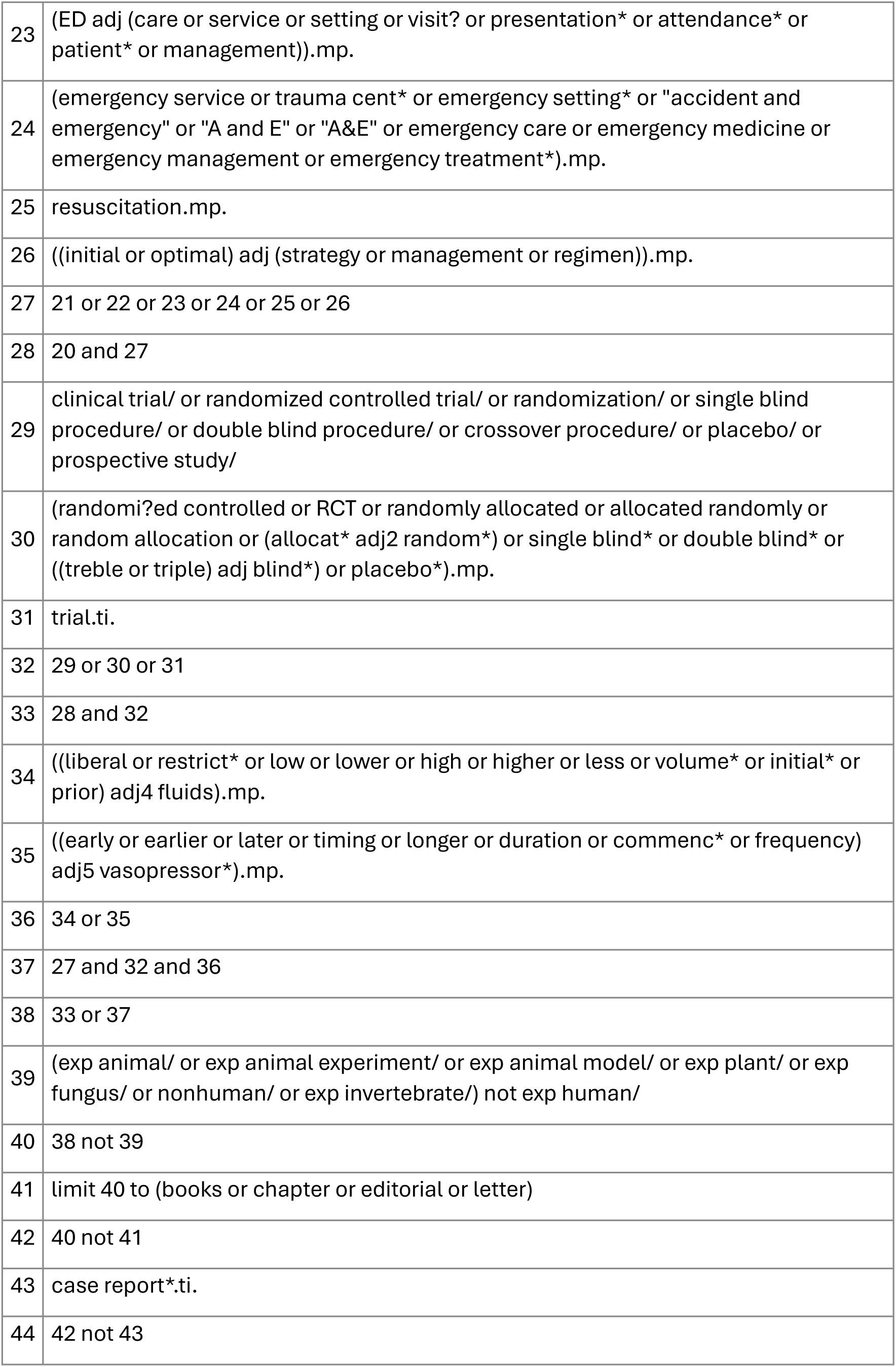

Database(s): **EBM Reviews - Cochrane Central Register of Controlled Trials**

Search Strategy:

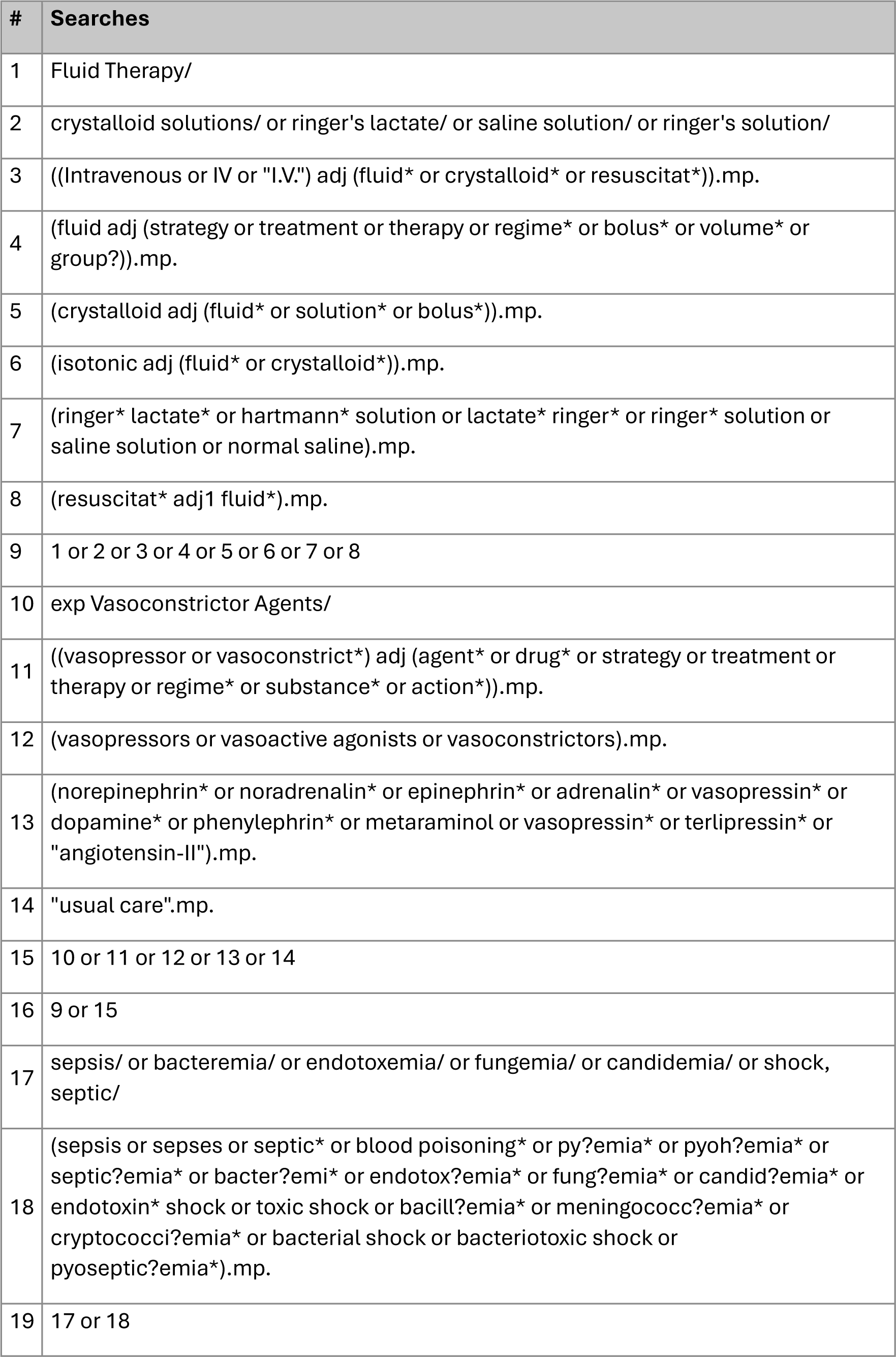
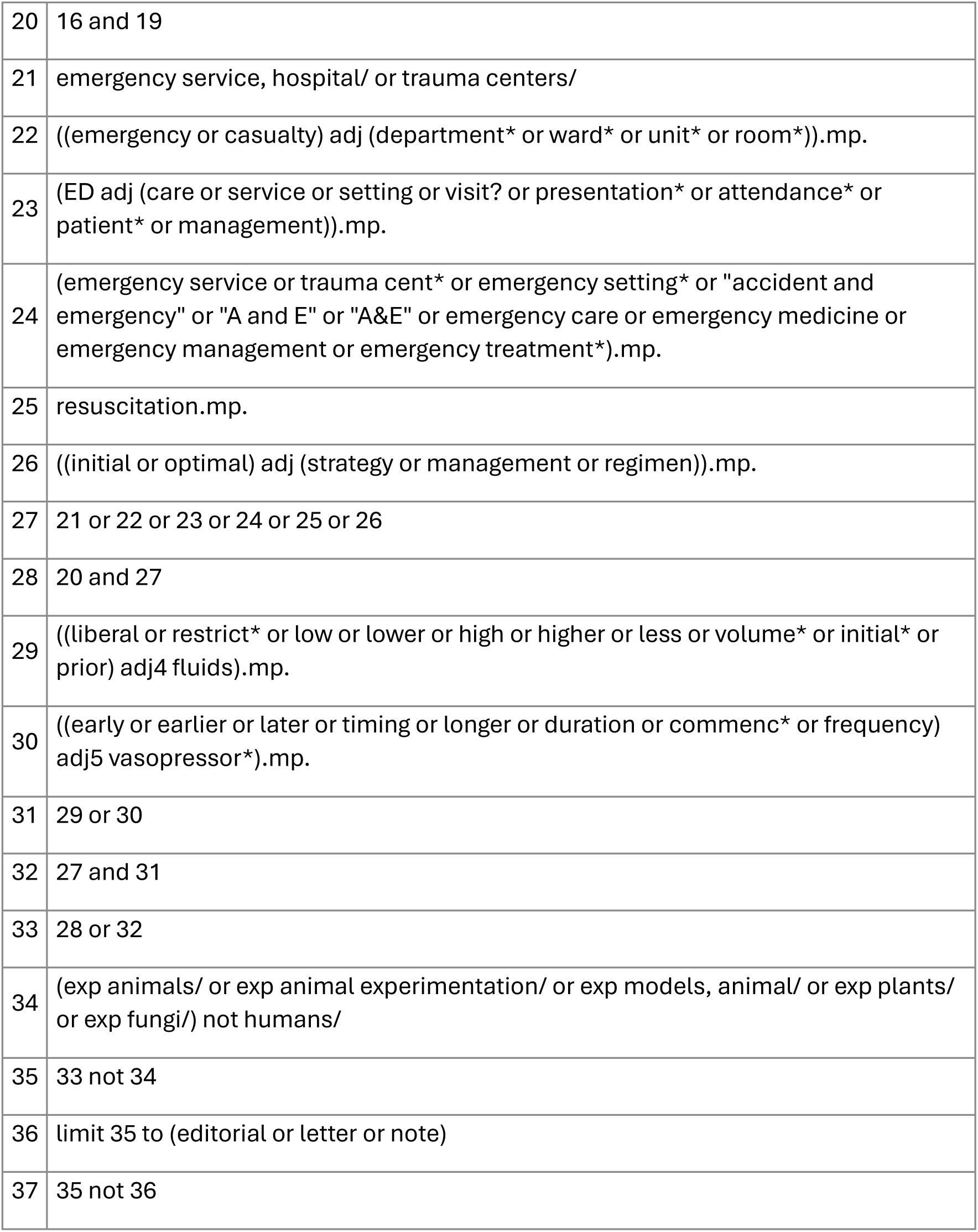

### WEB OF SCIENCE search strategy

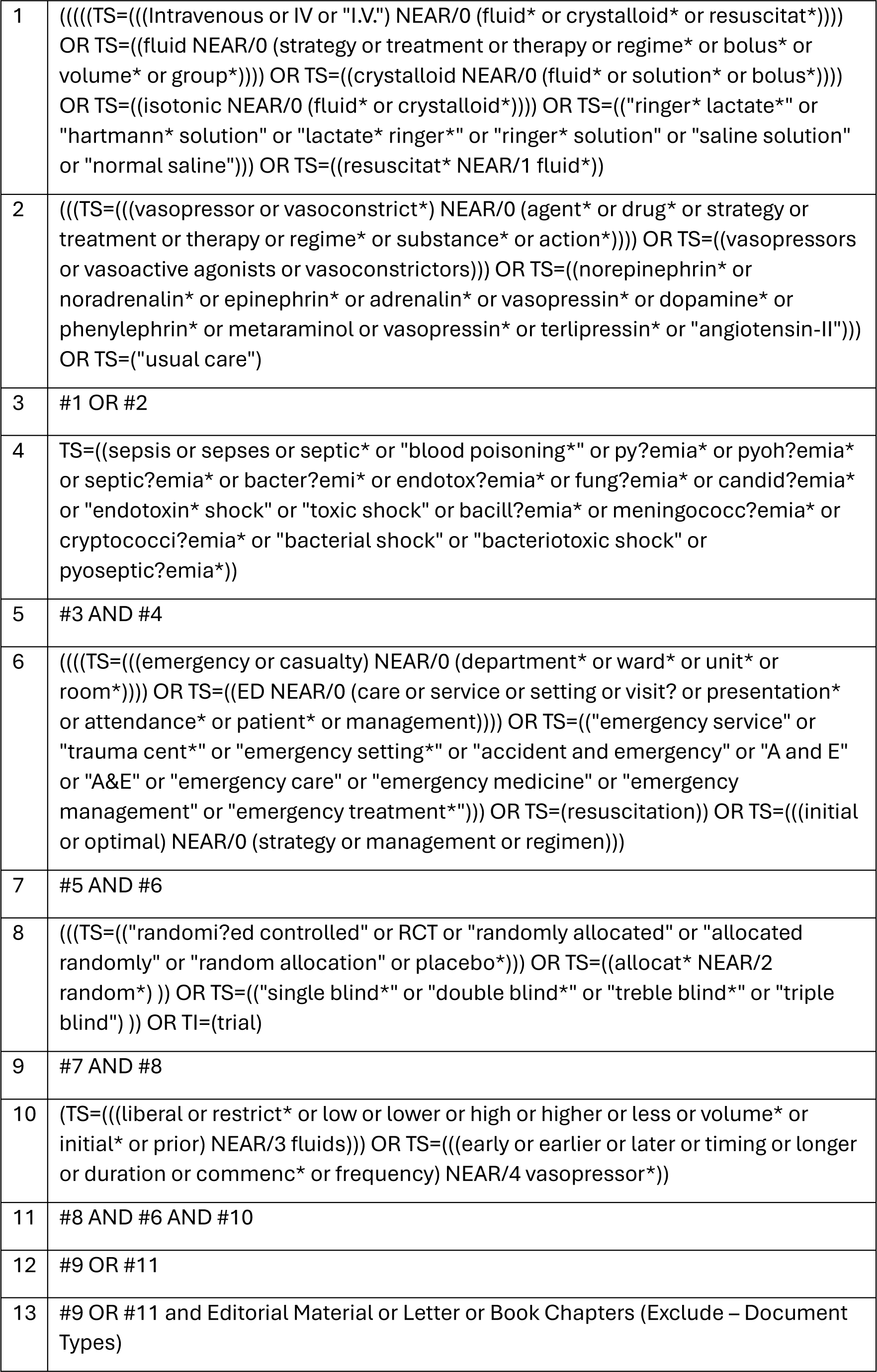

### Density Distribution of Prior Probabilities for Likely Effect

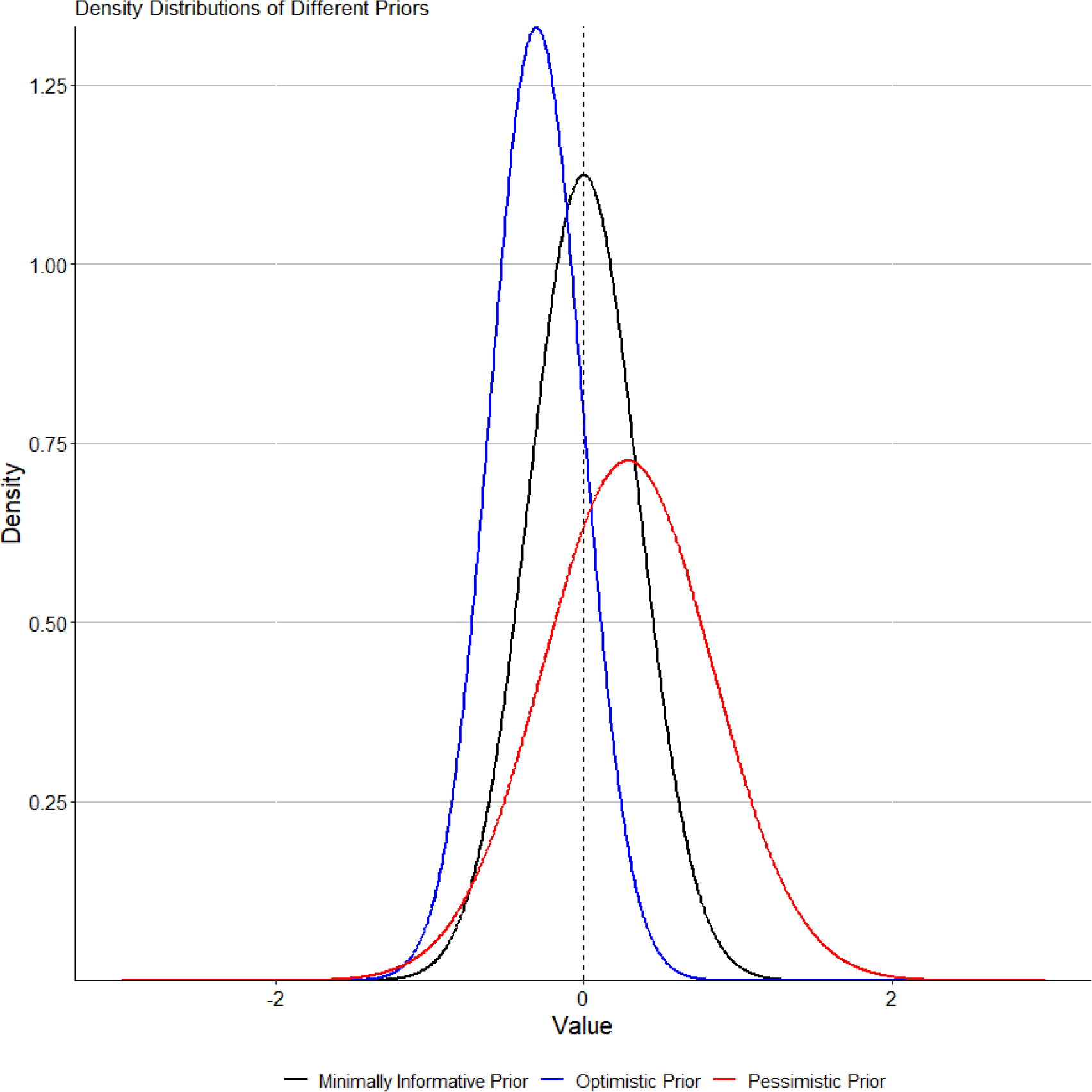

